# Genome Recombination between Delta and Alpha Variants of Severe Acute Respiratory Syndrome Coronavirus 2 (SARS-CoV-2)

**DOI:** 10.1101/2021.10.11.21264606

**Authors:** Tsuyoshi Sekizuka, Kentaro Itokawa, Masumichi Saito, Michitsugu Shimatani, Shutoku Matsuyama, Hideki Hasegawa, Tomoya Saito, Makoto Kuroda, COVID-19 Genomic Surveillance Network in Japan (COG-JP)

## Abstract

Prominent genomic recombination has been observed between the Delta and Alpha variants of severe acute respiratory syndrome coronavirus 2 (SARS-CoV-2) isolated from clinical specimens in Japan. It is necessary to intensively study such marked genetic variations and characterize the emerging variants after careful verification of their lineage and clade assignment.

## Text

### Ethics Statement

The study protocol was approved by the National Institute of Infectious Diseases, Japan (Approval no. 1091). The ethics committee waived the requirement for written consent with respect to research on viral genome sequences.

## Course of the Study

We conducted a genome surveillance of the severe acute respiratory syndrome coronavirus 2 (SARS-CoV-2) with help from the local public health centers, laboratories in research institutes, commercial laboratories (1, 2), and airport quarantine stations (3). The coronavirus disease-19 (COVID-19) Genomic Surveillance Network in Japan (COG-JP) has consistently monitored the prevalence of the Phylogenetic Assignment of Named Global Outbreak (PANGO) lineages of SARS-CoV-2 from the first COVID-19 case (January 15, 2020) to recent cases (September 30, 2021). Until now, 120,476 domestic and 2,018 quarantine isolates have been deposited in the Global Initiative on Sharing All Influenza Data (GISAID) EpiCoV database (1, 2). Whole-genome sequences were assigned to the SARS-CoV-2 isolates (≥ 29 kb genome size) obtained from domestic COVID-19-positive patients in Japan (n = 120,476), according to the PANGO lineage definition (v3.1.14, 2021-09-28) (4).

## Observations and Discussions

During the surveillance, we monitored several variants of concern (VOCs) of the PANGO lineage. We found six unique specimens (Fig. 1) among the 21A (Delta) clade isolates that exhibited a low quality assignment (“not determined” or “none”) in the PANGO lineage, even though their genome sequences had been completely determined with high read coverage throughout the whole genome region. A detailed genome alignment by Nextclade (5) suggested that despite being clonal isolates of the 21A (Delta) clade, these six isolates show identical mutation profiles with 20I (Alpha, V1), particularly between the *ORF6* and *N* genes, located towards the latter part of the SARS-CoV-2 genome (Fig. 1). Additionally, the alignment clearly revealed a possible recombination spot between the *ORF6* and *ORF7a* genes (Fig. 1). However, the next generation sequencing (NGS)-based read mapping analysis did not indicate the presence of any heterogeneous mix of alleles in the clinical specimens, suggesting that these patients had not acquired multiple variant infections with 21A (Delta) and 20I (Alpha, V1) clades at the acute stage of the infection (Fig. 2).

**Figure 1.**
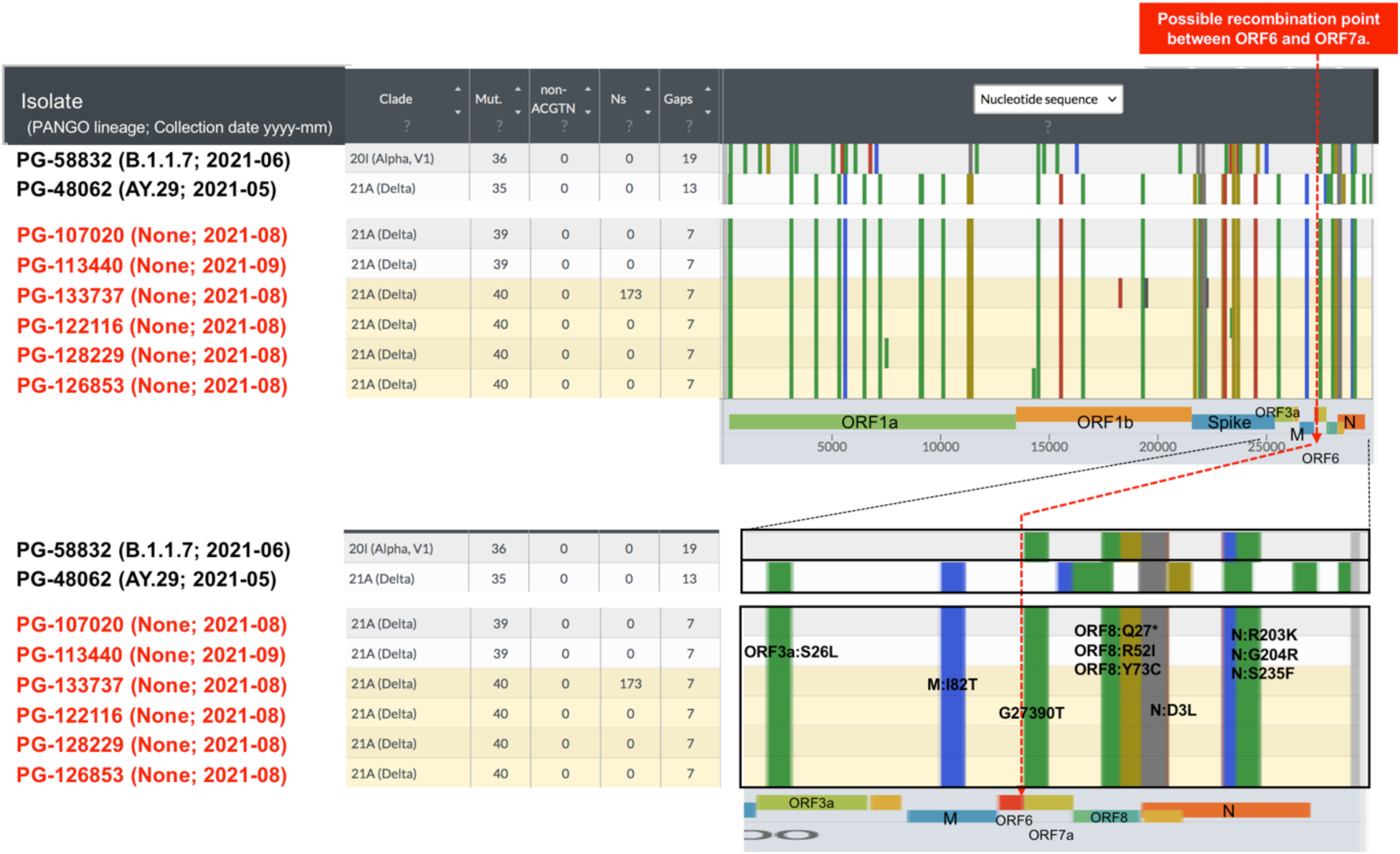
Clade assignment and pair-wise genome alignment of severe acute respiratory syndrome coronavirus 2 (SARS-CoV-2) samples were performed using Nextclade analysis (5). In a comparison with the control for B.1.1.7 Alpha variant (PG-58832) and B.1.617.2 Delta variant (PG-48062), Nextclade alignment showed that six isolates (highlighted in red) have been assigned in the 21A (Delta) clade, but their mutation profiles after the *ORF7a* gene suggest identical profiling with the B.1.1.7 Alpha variant (PG-58832). The possible recombination point is shown as red broken line.

**Figure 2.**
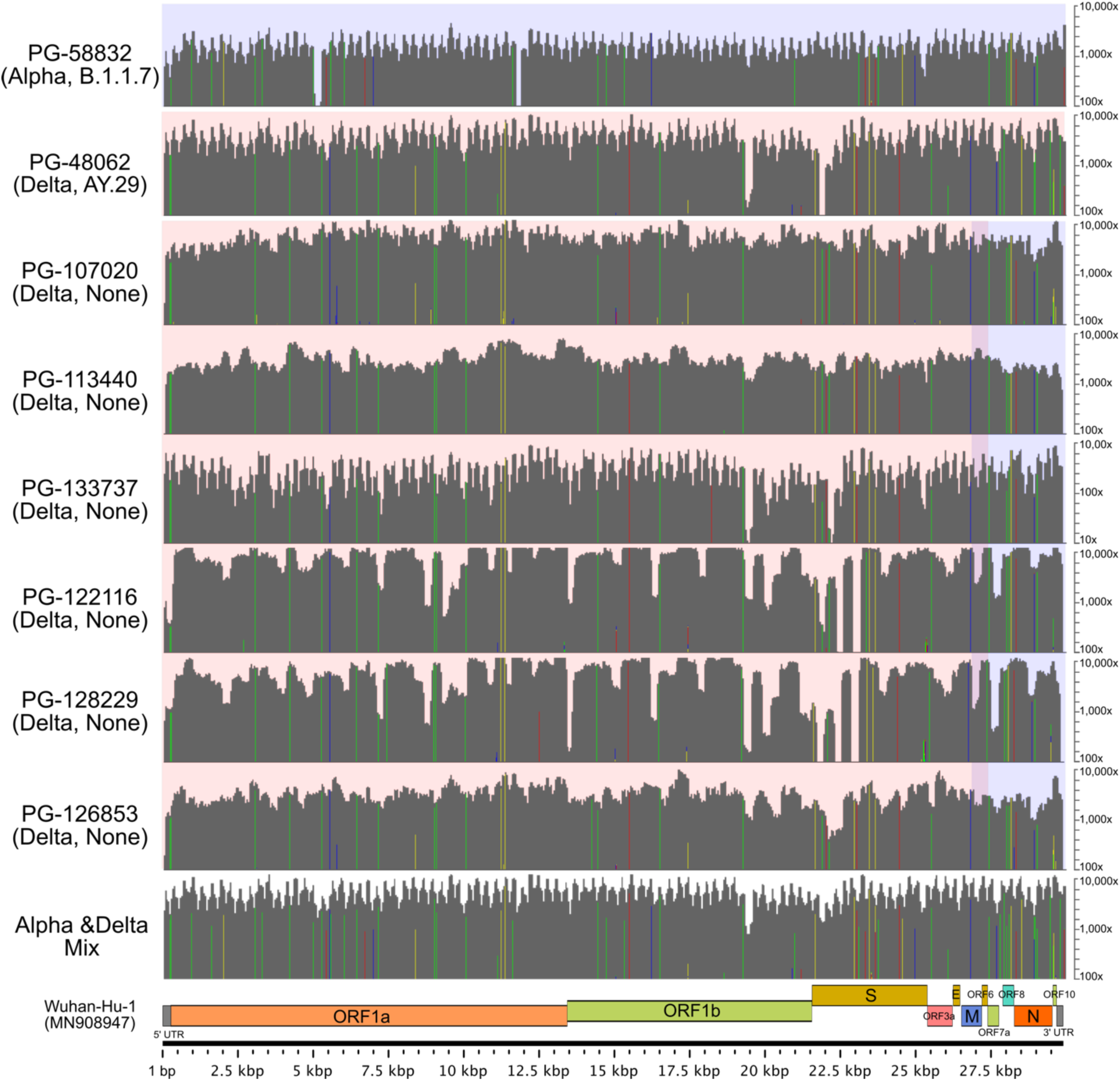
Read mapping analysis of the severe acute respiratory syndrome coronavirus 2 (SARS-CoV-2) recombination isolates. The new generation sequencing (NGS) reads were mapped to the whole genome sequence of Wuhan-Hu-1 (GenBank ID: MN908947), and the mutation profile of each isolate was compared using control isolates (PG-58832, Alpha B.1.1.7; PG-48062, Delta AY.29). A background color in dark blue and light red is indicated for the genome region corresponding to the Alpha and Delta variants, respectively. Detected nucleotide variations are highlighted with a vertical line (A: red, G: yellow, C: blue, and T: green) on the read mapping area in dark gray. Co-infection samples with Alpha and Delta variants were prepared virtually for comparison.

Interestingly, all six clinical isolates that exhibit the abovementioned recombination were detected around mid-August 2021. Even though the mutation profiles point towards the clonal nature of these six isolates (Fig. 1), there is no epidemiological link among the patients.

Incidentally, the two variants, namely 21A (Delta) and 20I (Alpha), were detected in 93% and 5% of the total specimens, respectively, in mid-August 2021 in Japan (6), suggesting the possibility of multi-variant infection in a single patient leading to such a recombination event. However, we have not yet identified a potential patient with mixed infection, i.e., one who has tested double positive in the variant polymerase chain reaction (PCR) test. Furthermore, the number of COVID-19-positive patients has been significantly decreasing in Japan since early October 2021. In parallel, such a recombinant isolate has not been detected thus far, indicating that it might be contained compared with other domestic Delta variants.

A recent report by Gribble et al. demonstrated that recombination is a critical event for creating the coronavirus diversity, and RNA proofreading exoribonuclease (nsp14-ExoN) is responsible for generating the recombination frequency as well as the altered recombination products in the in vitro culture experiments (7). The report has also described eight potential recombination hotspots at the microhomologous sequence of the SARS-CoV-2 genome. In fact, one of the hotspots, which lies between *ORF6* and *ORF7a*, could be the target site responsible for the recombination event observed in the current study. Interestingly, the recombination variant detected in this study carries a spike protein identical to the one in the domestic Delta variant, thereby suggesting that further risks would not be concerned with infectivity and immune escape.

The detection of SARS-CoV-2 variants with *ORF7a, ORF7b*, and *ORF8* deletions (8-11) can explain the occurrence of the abovementioned recombination event. In fact, the hotspots around *ORF7a* can facilitate the generation of a novel isolate that exhibits different genetic profiles owing to co-infection by distinct variants in the COVID-19 patient. In addition to the six isolates mentioned in this study, we also investigated the total deposits in GISAID (by 2021-09-30) for other possible genomic recombinations. We found a USA isolate [hCoV-19/USA/MO-CDC-LC0213262/2021 (EPI_ISL_4164992|2021-08-07)] portraying a recombination between the Delta and Alpha variants in *ORF7a*, but we could not reconfirm the validity of the event because we were unable to obtain the raw NGS sequencing reads for analysis.

## Conclusions

In conclusion, this is the first identification of a novel recombination SARS-CoV-2 variant between the 21A (Delta) and 20I (Alpha, V1) clades in domestic clinical specimens. As suggested by previous in vitro culture experiments (7), such a recombination can possibly be generated in the real world. Therefore, the simple PANGO and clade assignment might mis-identify notable variants based upon ordinary genome surveillance, and we must intensively monitor and carefully inspect such marked genetic variations to ensure their proper characterization.

## Data Availability

All data produced in the present study are available from GISAID and DNA Data Bank of Japan (DDBJ). Raw NGS sequencing reads have been deposited in the DNA Data Bank of Japan (DDBJ; accession number: DRA012825), as shown in the Table.　

## Acknowledgments

We would like to thank Rina Tanaka, Satsuki Eto, Risa Someno, Akina Ogamino, Naomi Nojiri, Hazuka Yoshida, Tomoko Ishihara, Tadaki Suzuki and Nozomi Takeshita for the whole-genome sequencing and data curation of the SARS-CoV-2 specimens. This work was supported by a Grant-in-Aid from the Japan Agency for Medical Research and the Development Research Program on Emerging and Re-emerging Infectious Diseases (JP21fk0108103) and a grant from the Ministry of Health, Labor, and Welfare, Japan (19HA1001 and 20HA2007). Finally, we would like to thank all the researchers who have deposited and shared genomic data of SARS-CoV-2 on GISAID.

## Disclaimers

The opinions expressed by the authors contributing to this article do not necessarily reflect the opinions of the Centers for Disease Control and Prevention or the institutions with which the authors are affiliated.

## Data availability

Raw NGS sequencing reads have been deposited in the DNA Data Bank of Japan (DDBJ; accession number: DRA012825), as shown in the Table.

**Table:** Summary information of the recombination variant isolates of SARS-CoV-2 ^1^

| Isolate | Sampling date<br>(yyyy-mm) | Clade | Lineage<br>(PANGOLIN <sup>2</sup>,<br>2021-09-28) | GISAID <sup>3</sup> ID | NGS <sup>4</sup> raw reads<br>(fastq) |
| --- | --- | --- | --- | --- | --- |
| PG-107020 | 2021-08 | 21A (Delta) | None | EPI_ISL_4882472 | DRR321268 |
| PG-113440 | 2021-09 | 21A (Delta) | None | EPI_ISL_4882473 | DRR321269 |
| PG-133737 | 2021-08 | 21A (Delta) | None | EPI_ISL_4882477 | DRR321273 |
| PG-122116 | 2021-08 | 21A (Delta) | None | EPI_ISL_4882474 | DRR321270 |
| PG-128229 | 2021-08 | 21A (Delta) | None | EPI_ISL_4882476 | DRR321272 |
| PG-126853 | 2021-08 | 21A (Delta) | None | EPI_ISL_4882475 | DRR321271 |

<sup>[1]</sup> SARS-CoV-2: Severe acute respiratory syndrome coronavirus 2
<sup>[2]</sup> PANGOLIN: Phylogenetic Assignment of Named Global Outbreak Lineages
<sup>[3]</sup> GISAID: Global Initiative on Sharing All Influenza Data
<sup>[4]</sup> NGS: Next generation sequencing

## Author Bio

Dr. Tsuyoshi Sekizuka is a chief at the National Institute of Infectious Diseases in Shinjuku-ku, Tokyo, Japan. His main research interest is pathogen genomics.

